# Physician Knowledge, Attitudes, and Practices Regarding Viral Exanthem Diagnosis And Mandatory Reporting Requirements in Major Metropolitan Regions of Florida, USA

**DOI:** 10.64898/2026.03.18.26348447

**Authors:** Winnie Chen, Sarah Ballarin, Maria Fioletova, Chanakya R. Bhosale, Thomas Matthews, Anna K. Potter, Jayson Forbes, Cyril Blavo

## Abstract

**Objective:** To assess physician knowledge, attitudes, and practices regarding viral exanthem diagnosis and mandatory reporting requirements among physicians practicing in metropolitan regions of Florida.

**Study Design:** An IRB-exempt cross-sectional survey was distributed via REDCap to practicing physicians and residents in family medicine, internal medicine, pediatrics, and infectious disease. The 19-item survey evaluated knowledge of viral exanthem diagnosis (measles, rubella, roseola), reporting requirements, physician attitudes, and clinical practices. Knowledge scores were compared by specialty using ANOVA with post-hoc testing. Multivariate analysis and linear regression examined relationships between physician confidence and knowledge performance.

**Results:** Of 162 respondents, 146 completed surveys were analyzed. Participants included pediatricians (n=74), family medicine physicians (n=48), and internal medicine physicians (n=24). The overall mean knowledge score was 78.5% (SD 20.5). Pediatricians achieved the highest scores (82.7%), significantly higher than family medicine physicians (73.3%; p=0.04). Differences in vignette-based diagnostic knowledge and mandatory reporting knowledge were not statistically significant across specialties. Roseola was the most commonly diagnosed viral exanthem (66%), followed by measles (30%) and rubella (17%). Most physicians (91.4%) reported interest in additional training.

**Conclusions:** Physicians demonstrated generally strong knowledge of viral exanthem diagnosis and reporting; however, gaps remain in clinical differentiation of similar rash presentations. Targeted physician education may improve diagnostic accuracy and public health reporting amid declining vaccination rates and increasing measles outbreaks.

**Funding Statement:** This study did not receive any funding.

## Introduction

Viral exanthems (VE) are rashes associated with systemic viral infections such as measles, rubella, and roseola [1]. They are often characterized by a diffuse eruption of macules and papules that can be erythematous in nature [2]. VE can be differentiated based on distinct patterns; however, definitive diagnosis cannot always be made solely on the rash’s appearance and symptoms. It is important for clinicians to incorporate a comprehensive patient history, including travel from endemic areas, vaccination records, prodromal symptoms, and potential exposure history, to aid diagnostic accuracy. While these infections are typically self-limiting, atypical or severe patterns can present in immunocompromised patients, children, and pregnant women [3]. Despite the availability of safe and tested vaccine options, VE, such as measles was the cause of 142,300 deaths worldwide, with the majority of cases found among young children and immunocompromised adults, driven by increased travel from endemic areas, limited vaccine availability in under-resourced areas and declining vaccination rates in regions where they were previously eliminated, indicating the continuing public health importance of these viruses [4, 5].

The measles virus is a paramyxovirus belonging to the genus *Morbillivirus* [6]. The measles virus is a highly contagious viral infection transmitted via respiratory or aerosolized droplets. It contains a CD150 (SLAMF1) receptor that allows it to infect immune cells of the respiratory tract, which then transports the virus to the lymphoid cells, further causing systemic disease and contributing to its high virulence and contagiousness [7]. After the virus gains entry, there is an incubation period of up to 3 weeks, which is followed by the prodromal phase of a high fever, often greater than 39 degrees Celsius [6]. Other early symptoms are cough, malaise, and the pathognomonic Koplik spots, which are small, white spots located on the buccal mucosa. The rash in measles begins on the face before spreading to the rest of the body. Due to overlapping symptoms with the common cold and other illnesses that have symptoms of a fever and rash, measles can be misdiagnosed [8]. Currently, the preferred method of diagnosing measles is NAAT. Serologic IgM can be used to make the diagnosis, but clinical caution should be exercised as patients within 72 hours of developing a rash can return false-negative serology up to 20% of the time [9]. Rubella virus is a non-vector-borne member of the family Togaviridae and the sole member of the genus, Rubivirus [10]. Rubella virus contains RNA, which is surrounded by a capsid and a lipoprotein envelope. Rubella is asymptomatic in 25% to 50% of young children with an incubation period of 2 to 3 weeks. Early symptoms are a mild fever, malaise, lymphadenopathy, and sore throat that precedes a pinpoint erythematous maculopapular rash. The classic rubella rash starts on the face and neck and proceeds to spread to the trunk and lower extremities over the course of 3 days [11]. Although this presentation is characteristic, the rubella virus can be asymptomatic in 25% to 50% of young children, with an incubation period of 2 to 3 weeks. Complications such as congenital rubella syndrome (CRS), which affect newborns when a pregnant woman is exposed to the rubella virus, manifest in the triad of cataracts, congenital defects, and deafness. Since clinical manifestations can range from mild to atypical presentations, clinical diagnosis is made not only on the symptoms, but also serological testing for rubella-specific IgM and/or IgG antibodies remains the gold standard of diagnosis [9].

Roseola infantum, also known as exanthema subitum or sixth disease, is predominantly caused by infection with human herpesvirus 6 (HHV-6), or, less commonly, by human herpesvirus 7 (HHV-7) [12]. About ninety percent of cases occur in children younger than two years. Its clinical course begins with a high fever of up to 40 degrees Celsius, lasting 3 to 5 days, followed classically by a maculopapular erythematous rash originating at the trunk as the fever resolves. A complication of roseola infantum is acute febrile seizures due to its ability to cross the blood-brain barrier. Roseola infantum is a clinical diagnosis without any need for serology. Unlike measles and rubella, roseola has no vaccine available, but it continues to affect young children predominantly, accounting for 10-45% of febrile illnesses in infants, warranting the need for proper public health measures to avoid spreading of the virus [12].

Although cases of VE, primarily measles, mumps, and rubella, have drastically decreased in the United States due to high vaccination rates, outbreaks and cases continue to occur in areas with high travel from endemic or under-resourced areas, including the State of Florida. These cases pose a public health threat in regions where proper public health measures, including vaccine coverage, continue to fall. While vaccine-preventable VE rates have plummeted following implementation of mandatory school vaccines in 1997 for middle school entry, Florida was the first state in the United States to move to end all mandates for vaccine requirements for children and adults on September 3rd, 2025, with planned removal of select vaccine mandates in January 2026 [13, 14, 15]. As of August 5th, 2025, there have been 1356 confirmed measles cases in 41 jurisdictions, with 32 outbreaks reported in 2025 compared to 16 outbreaks in 2024 [16]. Of the 1356 cases in 2025, 92% were unvaccinated or had unknown vaccine history, the majority under age 20 years old, with most of the cases being in Texas (800+) [https://www.cdc.gov/measles/data-research/index.html]. In 2024-2025, Florida had 30 confirmed cases of measles [17]. People are most protected through herd immunity when over 95% of the community is vaccinated; however, MMR vaccination coverage among kindergarteners in the USA has decreased to 92.7% in 2023-2024 from 95.2% during 2019-2020 [16]. In the United States, vaccines are readily available, but drivers of vaccine hesitancy include misinformation about side effects, reduced vaccination rates during and after the COVID-19 pandemic, health disparities among underserved groups in rural and urban settings, and a lack of awareness regarding the severity of infection, leading to potential public health risks [18]. The current and recent outbreaks of VE stress the importance of proper public health strategies to prevent future outbreaks, including continuing education of healthcare professionals, especially in primary care, pediatrics, and family medicine, where patients are more likely to present with complications of VE.

In light of current circumstances, it is increasingly critical for physicians and healthcare workers to remain vigilant in recognizing vaccine-preventable infectious diseases to aid in public health measures and avoid further outbreaks of these diseases. The purpose of this study is to determine the current knowledge, attitudes, and practices among practicing physicians regarding viral exanthem diagnosis and their mandatory reporting requirements in various metropolitan areas of Florida, USA. The specific objectives of our study are 1) to assess physicians’ ability to correctly identify and differentiate between clinically similar VE (rubella, measles, and roseola/HHV6) and their respective mandatory reporting requirements. 2) To evaluate physicians’ confidence in distinguishing these VEs and their views on the necessity of additional training. 3) To identify future steps regarding educational and public health interventions. We hypothesize that our data will reveal differences between physicians of different specialties or of different levels of exposure to these viruses.

## Methods

### Overall Study Design

This IRB-exempt cross-sectional study utilized an online 19-question survey measuring demographic information (e.g., specialty of medicine), knowledge, attitudes, and practices (KAP) among practicing health professionals in Florida. The methodology for this study was closely modeled after that of a preliminary study [19], with modifications to address the specific aims of the present investigation. See Appendix for the list of questions utilized **(Appendix 1)**. The inclusion criteria for this study were actively licensed physicians (DO or MD) or residents with a medical training license. Specialties of focus included primary care, family medicine/practice, pediatrics, internal medicine, and infectious disease. Overall, the questions tested participants’ knowledge of viral exanthem diagnosis, treatment, and risk factors. No intervention or treatment was used in this study.

### Participants and Specialties

Prior research has established deficiencies in knowledge of VE amongst physicians. However, no studies specifically measured responses from physicians practicing in the state of Florida, and no studies have examined mandatory reporting requirements in the United States. Potential participants were first identified utilizing the Florida Department of Health public practitioner profile database of healthcare professionals, filtered by license in “active” status as of November 2024, by county, and specialty of choice, with a cap of 1050 practitioners per specialty from each county. The specialties of choice were family medicine, internal medicine, pediatrics, and infectious disease. If the database for a specific county listed fewer than 200 professionals meeting these criteria, all practitioners of that specialty were included. A broad county-based search strategy was used. Emails of n=1085 family medicine, n=1265 pediatric, and n=491 infectious disease doctors were collected.

### Survey Instrument

In order to ensure the anonymity and safety of survey responses, the secure REDCap survey platform was utilized to distribute our survey and capture data. REDCAP facilitated sending survey invites, retrieving responses, and initiating data analyses. Surveys were sent out randomly, on a weekly basis, and this was repeated up to ten times if no response was received. The survey consisted of six vignette-style knowledge questions; three questions that addressed diagnosing a viral exanthem, and the other three questions focused on mandatory reporting of VE. There were five attitude questions: 1. regarding the clinical process of diagnosis, 2. physician training on diagnosing VE, 3. misdiagnosis/underdiagnosis of VE, 4. Confidence in diagnosing VE, and 5. confidence of properly reporting VE. Two practice questions assessed 1. perceived childhood vaccination rates among respective patient populations, and 2. what VE physicians have diagnosed.

### Data Analysis

For each question type, a sample proportion (%) was calculated by dividing the number of positive responses by the total number of survey responses.

### Specific Statistical Analysis

Survey responses regarding practicing specialty were consolidated into either family medicine, internal medicine, or pediatrics. Infectious diseases and other subspecialties of internal medicine were sorted under internal medicine unless otherwise specified by the surveyor. Analysis of variance (ANOVA) was conducted in the comparison of knowledge and attitude questions by specialty. This was followed by pairwise comparisons to determine significance between groups. A multivariate analysis of variance was used to compare vignette-style diagnosis and mandatory reporting knowledge questions. The relationships between vignette-style diagnosis knowledge questions and confidence attitude questions were analyzed through linear regression. Results of the complete statistical analysis are given in Tables #2-6.

### IRB & Human Subjects Disclaimer

The research was reviewed and was exempted by the Institutional Review Board of the Dr. Kiran C. Patel College of Osteopathic Medicine at Nova Southeastern University (NSU-KPCOM) on 12/20/2024. IRB number: 2024-655. The questionnaire included a prompt to receive informed consent electronically.

## Results

### Demographics

A total of 162 practicing physicians across Florida completed the survey. Among these, 146 participants provided complete data, representing various specialties such as family medicine (FM) (n=48), internal medicine (IM) (n=24), and pediatrics (n=74). Related subspecialties (n=16) were included in the overall descriptive demographic reporting, but were excluded from inferential analysis of knowledge and attitude-based questions. Of 162 responses, reported durations of practice among participating physicians spanned from less than 1 year to 53 years. The mean number of years in practice was 21.96 years with a standard deviation (SD) of 13.72. The median and mode number of years in practice was 20 years. Represented training backgrounds of participants included MD (n=118), DO (n=40), MD resident (n=3), and DO resident (n=1) (**Table 1**).

**Table 1.** Demographics table for the physician responses.

| Characteristics | # of Physicians | % of Physicians |
| --- | --- | --- |
| Years Practicing |  |  |
| 5 and under | 29 | 18% |
| 5–10 | 13 | 8% |
| 11–20 | 41 | 25% |
| 21–30 | 39 | 24% |
| 31–40 | 17 | 10% |
| 40 and over | 23 | 14% |
| License |  |  |
| MD | 118 | 73% |
| DO | 40 | 25% |
| Resident | 4 | 2% |
| Region |  |  |
| South FL | 66 | 28% |
| Central FL | 44 | 19% |
| North FL | 36 | 16% |
| Not reported | 16 | 7% |
| Specialty |  |  |
| Pediatrics | 73 | 45% |
| Family Medicine (FM) | 44 | 27% |
| Internal Medicine (IM) | 28 | 17% |
| Pediatrics, IM | 4 | 2% |
| Not reported | 13 | 8% |
| Location |  |  |
| Urban | 67 | 41% |
| Suburban | 64 | 40% |
| Rural | 12 | 7% |
| Urban, Suburban | 13 | 8% |
| Suburban, Rural | 3 | 2% |
| Urban, Suburban, Rural | 3 | 2% |

A total of 19 counties, out of 140 that provided responses, were represented based on participants’ self-reported practice locations. The counties, listed from most to least represented, were: Miami-Dade (n=30), Duval (n=19), Broward (n=18), Hillsborough (n=18), Orange (n=17), Palm Beach (n=9), Escambia (n=5), Leon (n=4), St. Johns (n=3), Pinellas (n=3), Lee (n=3), Volusia (n=2), Polk (n=2), Marion (n=2), St. Lucie (n=1), Seminole (n=1), Pasco (n=1), Collier (n=1), and Clay (n=1) (**Figure 1**). Of 162 responses, geographic practice settings reported by participants included rural (n=7), urban (n=97), and suburban (n=71) (**Table 1**). Business practice settings included academic (n=53), community hospital or clinic (n=51), private practice (n=60), telemedicine (n=9), and other (n=12). Participants were permitted to select multiple categories for both setting questions to most accurately reflect physicians’ practice environments.

**Figure 1.**
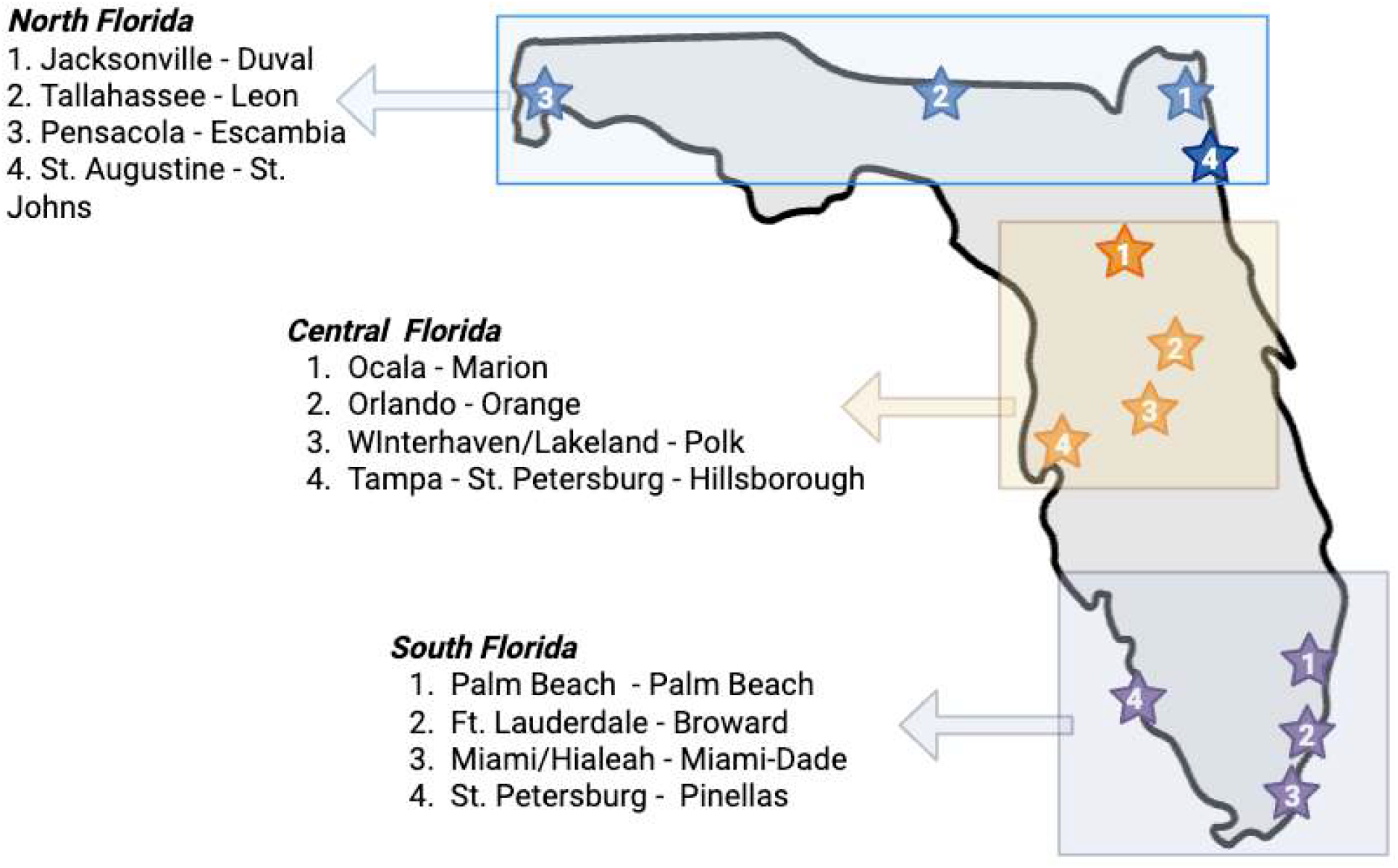
Map of Florida with major urban areas targeted in the study. i.e. More South Florida counties were targeted to encompass the South Florida metropolitan area. Map created in Biorender.com with license to Chanakya R. Bhosale.

### Overall Knowledge Performance

Across 146 participants, overall knowledge scores for vignette and mandatory reporting questions ranged from 16.67% to 100% with a mean of 78.54% (SD = 20.53). Specialty-specific knowledge means (M) showed that pediatricians (n = 74) scored the highest (M = 82.66%, SD = 16.42), followed by internal medicine physicians (n = 24) (M = 76.39%, SD = 25.02), and family medicine physicians (n = 48) (M = 73.26%, SD = 22.74) (**Table 2**). A one-way analysis of variance (ANOVA) demonstrated that differences in overall knowledge scores between specialties were significant (F(2,143) = 3.31, p = 0.04) (**Table 4**). Furthermore, the Tukey post-hoc comparisons revealed that only the pediatricians group scored significantly higher than family medicine physicians by 9.39% (p = 0.04) (**Table 3**). Whereas there were no significant differences between pediatrics and internal medicine, or between family medicine and internal medicine.

**Table 2.** Overall Knowledge Scores by Specialty.

| Specialty | n | Mean (%) | SD (%) | Min (%) | Max (%) |
| --- | --- | --- | --- | --- | --- |
| Family Medicine | 48 | 73.26 | 22.74 | 16.67 | 100 |
| Internal Medicine | 24 | 76.39 | 25.02 | 16.67 | 100 |
| Pediatrics | 74 | 82.66 | 16.42 | 33.33 | 100 |
| **Total** | 146 | 78.54 | 20.53 | 16.67 | 100 |

**Table 3.** Vignette and Mandatory Reporting Specific Knowledge Scores by Specialty.

| Specialty | n | Vignette Knowledge<br>Mean (%) | SD (%) | Mandatory<br>Reporting Mean (%) | SD (%) |
| --- | --- | --- | --- | --- | --- |
| Family Medicine | 48 | 65.0<sup>1</sup> | 27.1<sup>1</sup> | 78.3<sup>2</sup> | 23.0<sup>2</sup> |
| Internal Medicine | 24 | 72.6<sup>1</sup> | 28.0<sup>1</sup> | 79.7<sup>2</sup> | 25.1<sup>2</sup> |
| Pediatrics | 74 | 75.2<sup>1</sup> | 27.2<sup>1</sup> | 89.3<sup>2</sup> | 16.4<sup>2</sup> |
| **Total** | 146 | 71.4 | 27.5 | 84.1 | 20.9 |

<sup>1</sup> Vignette-based knowledge questions assessing the ability to differentiate measles, rubella, and roseola.
<sup>2</sup> Mandatory reporting knowledge questions assessing understanding of reporting requirements.

**Table 4.**
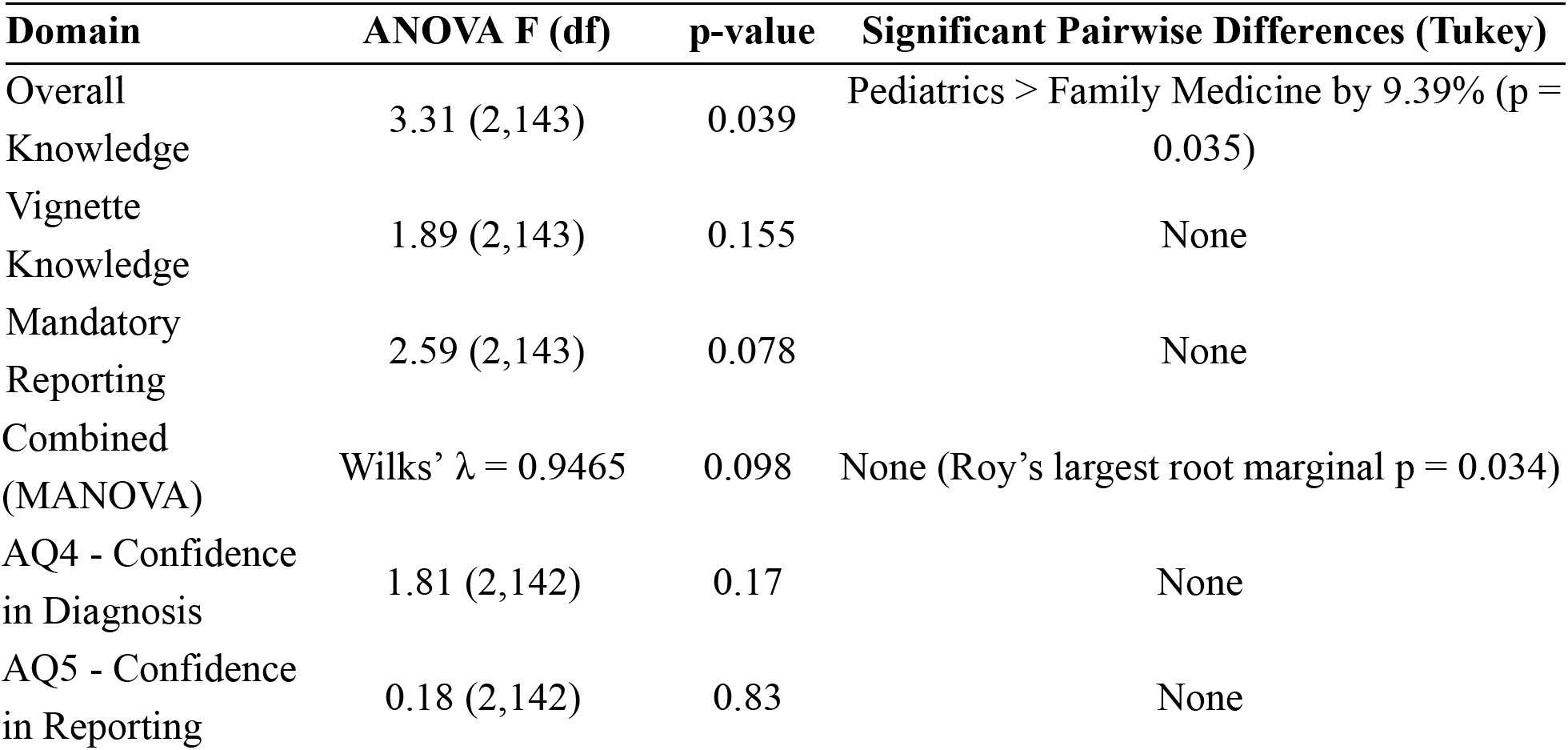
ANOVA Results.

| Domain | ANOVA F (df) | p-value | Significant Pairwise Differences (Tukey) |
| --- | --- | --- | --- |
| Overall Knowledge | 3.31 (2,143) | 0.039 | Pediatrics > Family Medicine by 9.39% ($p = 0.035$) |
| Vignette Knowledge | 1.89 (2,143) | 0.155 | None |
| Mandatory Reporting | 2.59 (2,143) | 0.078 | None |

|  |  |  |  |  |
| --- | --- | --- | --- | --- |
| Combined<br>(MANOVA) | Wilks' $\lambda = 0.9465$ | 0.098 | None (Roy's largest root marginal $p = 0.034$) | |
| AQ4 - Confidence<br>in Diagnosis | 1.81 (2,142) | 0.17 | None |  |
| AQ5 - Confidence<br>in Reporting | 0.18 (2,142) | 0.83 | None |  |

### Vignette-based Knowledge Questions

For vignette-based knowledge questions that assessed physicians’ ability to differentiate between measles, rubella, and roseola, pediatrics had the highest score. However, the ANOVA results and Tukey post-hoc comparisons revealed no significant difference between specialties when vignette-based knowledge questions were examined separately (F(2,143) = 1.89, p = 0.15) (**Table 3, Table 4**).

### Mandatory Reporting Knowledge Questions

Similarly, when knowledge questions about mandatory reporting requirements were examined separately, results showed no significant differences among the three specialties. Although overall ANOVA approached significance (F(2,143) = 2.59, p = 0.08), the Tukey pairwise comparisons were not significant (**Table 3, Table 4**).

To evaluate whether specialty had a combined effect on the diagnosis of VE and mandatory reporting knowledge, a multivariate analysis of variance (MANOVA) was conducted. Results demonstrated no statistically significant difference as indicated by Wilks’ lambda (lambda = 0.10, F = 1.98, p = 0.10), Pillai’s trace (p = 0.10), Lawley-Hotelling trace (p = 0.10), and Roy’s largest root (p = 0.03) (**Table 4**). While Roy’s largest root was marginally significant, this result can not be supported by the other indices, which showed no significant difference.

### Global Physician Attitudes

Physician opinions on the complexity and time burden associated with diagnosing VE (AQ2) were relatively evenly distributed, with 56.17% of respondents indicating that the diagnostic workup of VE was complex and time-consuming, whereas 43.83% indicated otherwise (**Figure 1**). Regarding interest in increased training on the workup and management of VE (AQ3), a substantial majority (91.36%) expressed a desire for additional training. This finding is consistent with perceptions of diagnostic inadequacy of VE (AQ1), where 26.54% of respondents felt that VE were certainly underdiagnosed or misdiagnosed, while 0.62% believed they were not.

**Figure 1.**
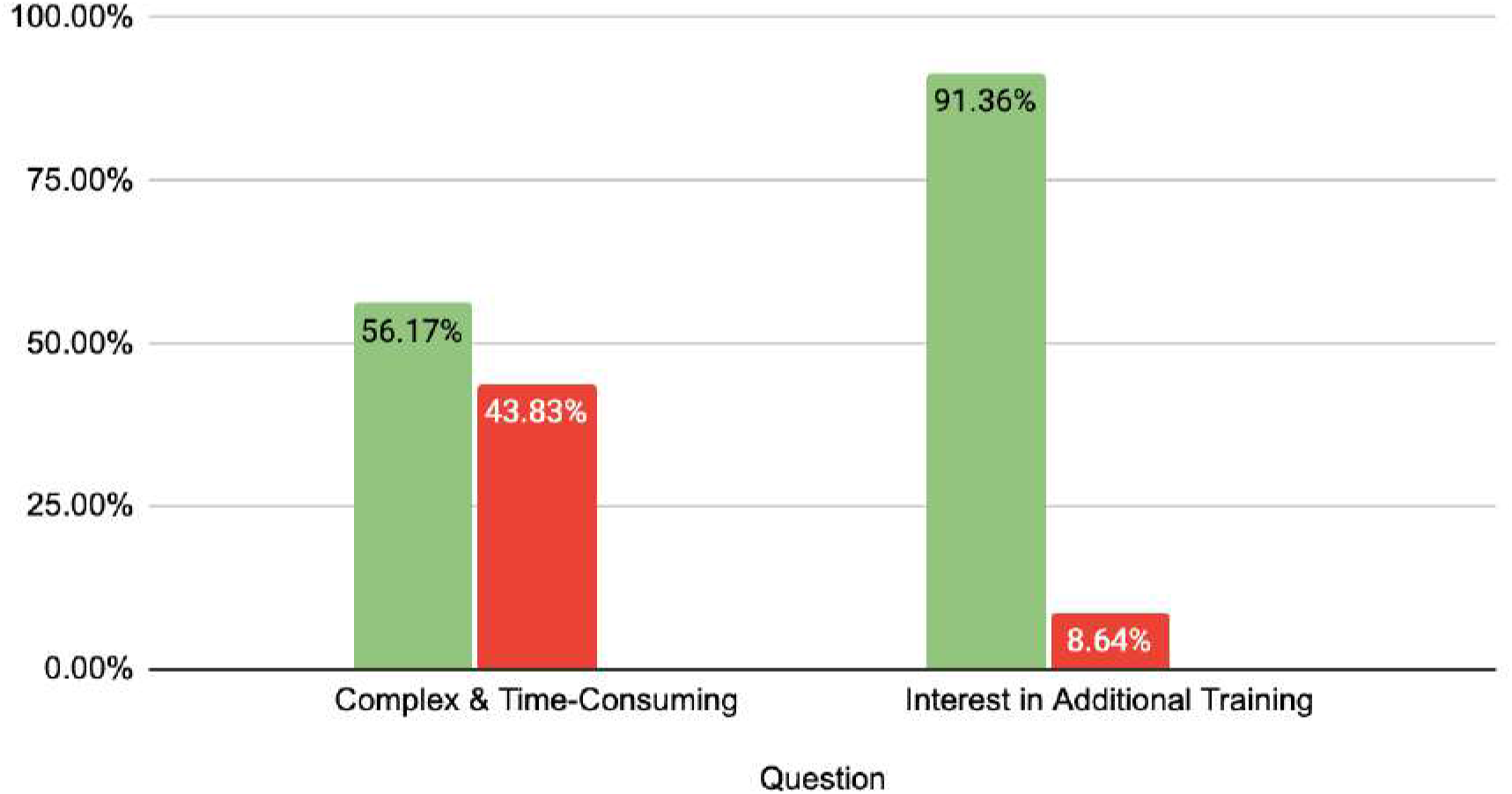
Global Physician Attitudes responses (green color indicating answer “yes” and red color indicating answer “no”).

### Specialty-based Differences in Attitudes

Physicians’ confidence in diagnosing VE (AQ4) and properly reporting a diagnosis of rubella, varicella, measles, and roseola to public health authorities was evaluated for specialty-based differences in attitudes (AQ5). The ANOVA for AQ4 yielded F(2,142) = 1.81, p = 0.17 and F(2,142) = 0.18, p = 0.83 for AQ5 (**Table 4**). These results showed that neither attitude question had a significant difference between specialties. Additionally, post-hoc comparisons were not significant.

### Associations between Knowledge Scores and Physician Attitudes

Two linear regression models were used to evaluate the relationship between knowledge scores and physician attitudes. The first linear regression model evaluated whether physicians’ confidence in diagnosing VE predicted vignette-based knowledge questions (AQ4). Results showed a significant positive relationship with every one-point increase in positive attitude, vignette-based knowledge scores increased by 10.73% (F(1,143) = 12.93, p < 0.001) (**Table 5**).

The second linear regression model examined the relationship between physicians’ perceived confidence in mandatory reporting requirements and mandatory reporting knowledge scores. Results also showed a significant positive relationship with every one-point increase in positive attitude, mandatory reporting knowledge scores increased 9.68% (F(1,143) = 20.40, p < 0.001) (**Table 5**).

**Table 5.** Linear Regression of Physician Attitudes Predicting Knowledge Scores.

| Outcome | Predictor | $\beta$ (%) | F (df) | p-value |
| --- | --- | --- | --- | --- |
| Vignette-based<br>knowledge score | Confidence in diagnosing viral<br>exanthems (AQ4) | 10.73 | 12.93 (1,143) | <0.001 |
| Mandatory reporting<br>knowledge score | Confidence in mandatory reporting<br>requirements (AQ5) | 9.68 | 20.40 (1,143) | <0.001 |

**Table 6.**
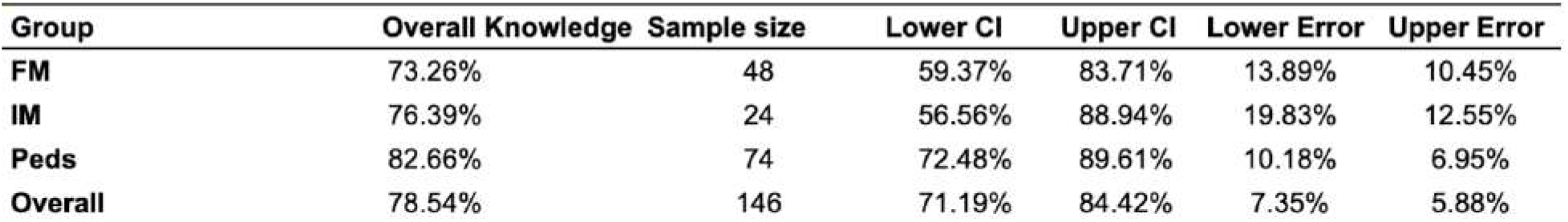
Wilson Confidence Intervals.

The responses from physicians who diagnosed measles, rubella, and roseola during their career was evaluated. It was found that roseola was the most frequently diagnosed VE, with a 66% response rate. Following roseola, measles had a 30% response rate, and rubella had a 17% response rate (**Figure 2**).

**Figure 2.**
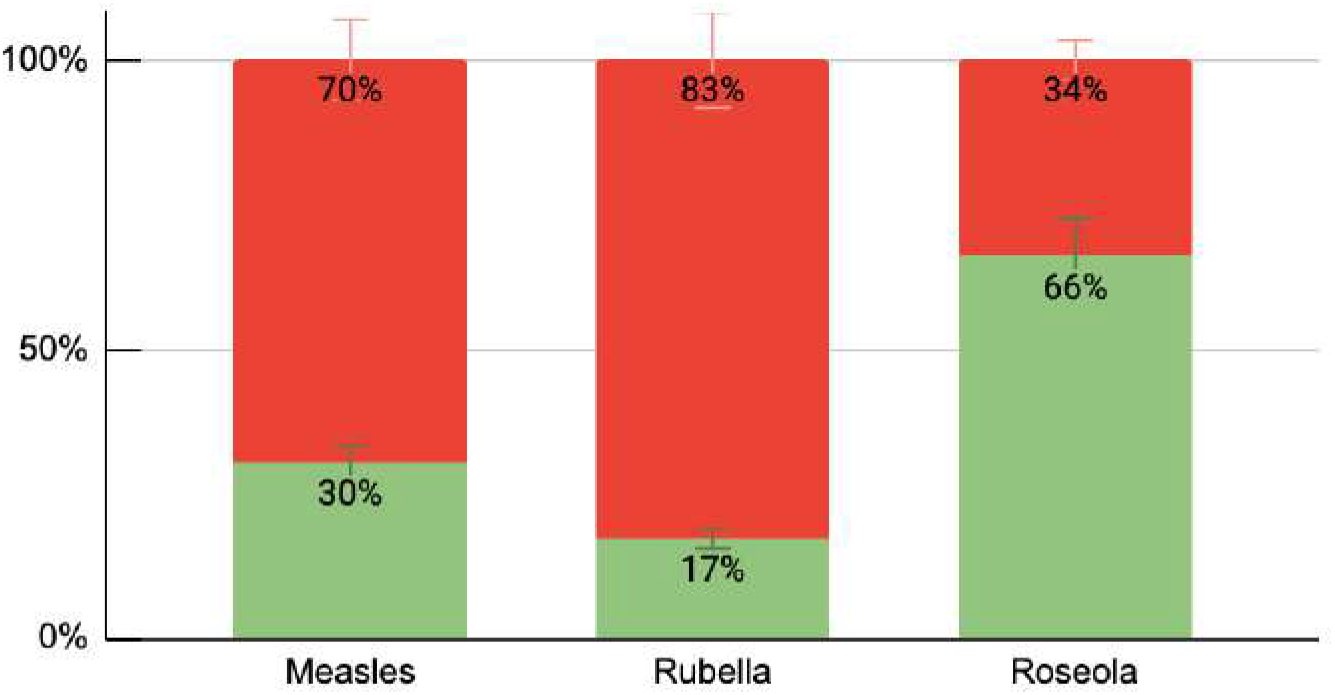
Percentages of physicians who diagnosed measles, rubella, and roseola during their practice. The green bar indicates physicians answering questions correctly, and the red represents incorrect responses.

Furthermore, perceived vaccination rates were evaluated based on physicians’ responses. 52.47% of physicians stated that less than 10% of children were vaccinated in their area of practice, 18.52% stated that 10-20% of children were vaccinated, 3.09% stated that 21-30% children were vaccinated, and 4.32% stated that more than 30% of children were vaccinated (**Figure 3**).

**Figure 3.**
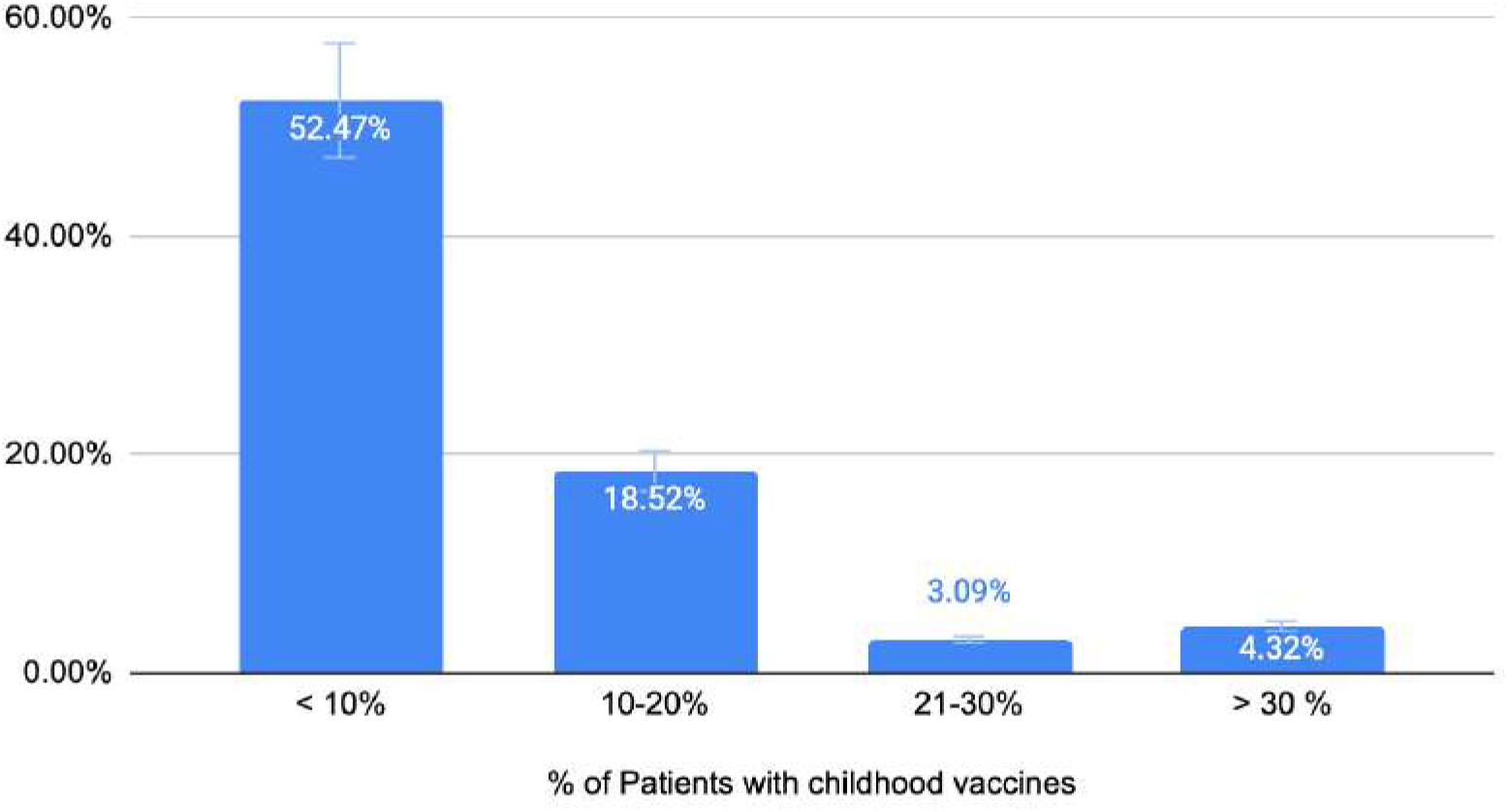
Perceived Vaccination Rates

## Discussion

In this survey, physicians practicing in Florida were assessed for their knowledge, attitudes, and opinions related to the diagnostic workup of VE, and clinical practices pertaining to VE (measles, rubella, roseola). Physician responses across these domains revealed knowledge gaps and limited confidence in the diagnosis and mandatory reporting of VE. Furthermore, our study confirms that Florida physicians have diagnosed rubeola, measles, and rubella in the state, affirming the continuing and emerging public health importance of VE in this region of the Southeastern United States.

The knowledge-based question on which physicians demonstrated the lowest performance involved diagnosing rubella from a clinical vignette, whereas the highest-scoring question addressed mandatory reporting guidelines of rubella. These findings suggest that physicians may be familiar with reporting once viral exanthem diagnoses are made, but clinical differentiation of similar rash presentations remains a challenge. Among specialties, pediatricians achieved the highest mean score, followed by IM and FM. This distribution is consistent with the epidemiology of VE, which is commonly present in the pediatric population [20]. Mean knowledge scores were statistically comparable among specialties, with the exception of pediatricians who had a statistically significantly higher mean score than FM physicians. This finding was somewhat unexpected, as ambulatory pediatricians and FM physicians are typically the first to encounter VE cases, while involvement of an IM or ID physician more often requires a referral or for the patient to first be seen by an emergency medicine physician.

Although specialty-based differences in vignette-based and mandatory reporting knowledge were not statistically significant when examined independently, there was an overall trend toward lower performance among non-pediatric specialties. This suggests that experience and clinical exposure may influence applied knowledge. This could be attributed to multiple reasons. One reason is that pediatricians and infectious disease physicians have more exposure to VE and thus must be constantly up-to-date on regulations and mandatory reporting. Importantly, multivariate analyses did not demonstrate a combined specialty effect on diagnostic and reporting knowledge, indicating that gaps are present across disciplines rather than limited to a single specialty.

Findings on physician attitudes further contextualize these findings. The majority of respondents reported feeling “somewhat confident” in diagnosing and reporting VE. There was a significant positive correlation found in self-reported confidence levels and knowledge scores, which suggests that confidence accurately reflects physician knowledge rather than being a result of overconfidence. These findings, in conjunction with the near-unanimous physician desire for increased training in addressing and managing VE in Florida underscores perceived knowledge deficits and discrepancies. Structured educational reinforcement could bridge knowledge gaps and restore physician confidence. Additionally, incorporating refresher modules or didactic content within continuing medical education may meaningfully improve outcomes and mandatory reporting practices.

The trend of declining vaccination rates has been an ongoing topic of concern for years. Vaccination coverage has been experiencing a trending decline in Florida among kindergartners since the 2018-2019 school year, with coverage for school-mandated vaccines dropping below 90% in the 2024-2025 school year, undermining herd immunity [21]. Our study shows that, correspondingly, some vaccine-preventable diseases have seen an increase in the United States in recent years. The most memorable instance has been the recent surge of measles cases across the country, predominantly in areas with lower vaccination rates [16]. Also notable has been the increase in pertussis in 2024 and 2025, with reported cases in 2024 being 6 times higher than pre-pandemic 2019 rates [22]. Other vaccine-preventable diseases, including mumps, toxigenic *C. diphtheria*, rubella, invasive pneumococcal disease, and type B *H. influenza*-associated epiglottitis, have not reported significantly increased national rates, which may be attributed to high overall vaccination rates [23, 24, 25, 26, 27]. These trends are a warning to practicing health professionals across the country and possibly threaten major setbacks for public health. Due to vaccine mandates, newer generations of Florida physicians who began practicing in this century have encountered the lowest number of cases of vaccine-preventable disease in United States history, leading to concerns regarding lack of experience in recognition and management of these diseases [28, 29].

### Limitations

This study has potential limitations that should be considered. These limitations are primarily methodological. First, the process of participant recruitment relied on email addresses provided publicly by the Florida Department of Health practitioner database, with a direct focus on FM, pediatrics, and infectious disease physicians. IM physicians were not initially targeted during the email collection process. Instead, they were identified through subspecialty categorization of existing survey responses. This represents a sampling limitation that may have contributed to uneven specialty representation and response rates. Secondly, the study focused mainly on FM, pediatrics, and IM, which may not be fully representative of the knowledge and practices of VE across all specialties, such as emergency medicine, dermatology, and obstetrics, where VE may also be encountered. Therefore, the study’s findings are more reflective of primary-care-based settings. Responses were also obtained from multiple metropolitan regions across Florida. This may result in the overall response rate being limited, and some counties were minimally represented. This geographic imbalance may limit the generalizability of findings to all practice settings, particularly in rural or underserved areas.

### Future Directions

Future research should explore physician-reported barriers to VE diagnosis and mandatory reporting guidelines to identify areas for intervention, which may include structured continuing medical education modules on the clinical differentiation of exanthems with similar clinical presentations. The same applies to a broader, national version of this study with a larger sample size and a more robust dataset. A Florida county-based analysis using mis/underdiagnosis rates of VE could aid in identifying high-risk areas in particular need of targeted public health and healthcare outreach to improve awareness, early recognition, and management of VE. A similar analysis could be conducted by analyzing the correlation between VE knowledge in rural and urban practice settings. Additionally, it would be interesting to see whether there is any association between years of practice and accuracy of VE diagnosis, particularly for physicians in practice prior to the adoption of mandatory school vaccines by Florida.

## Conclusion

The study evaluates physician knowledge, practice, and attitudes regarding viral exanthem diagnosis and mandatory reporting requirements in Florida. Physicians demonstrated high knowledge and understanding in most cases, with pediatricians scoring significantly higher than family doctors. There was a strong statistical correlation between knowledge scores and physician confidence in diagnosis, underlining the importance of additional training for more accurate diagnosis and comprehensive management. Additionally, knowledge gaps were identified, particularly in the clinical differentiation of rubella. These knowledge gaps are present in all specialties, demonstrating a systemic nature rather than confinement to a single discipline.

This topic is currently clinically relevant, as declining vaccination rates and increasing measles outbreaks carry important public health implications. Therefore, early diagnosis and reporting of VE are critical for outbreak prevention and control. Future studies are necessary on a larger scale, in different geographical areas, and among other physician specialties to evaluate differences, identify shared patterns, and address knowledge gaps in viral exanthem diagnosis.

## Data Availability

All data produced in the present work are contained in the manuscript.

## Acknowledgements

The authors would like to thank all participants who generously contributed their time and effort to complete the surveys for this study. Their participation was essential to the success of this research. The authors also acknowledge the Florida Department of Health for providing a publicly accessible platform that facilitated survey distribution and data collection.

## Notes

### Competing Interest Statement

The authors have declared no competing interest.

### Author Declarations

The research was reviewed and was exempted by the Institutional Review Board of the Dr. Kiran C. Patel College of Osteopathic Medicine at Nova Southeastern University (NSU-KPCOM) on 12/20/2024. IRB number: 2024-655.

